# Sudden hearing loss following vaccination against COVID-19

**DOI:** 10.1101/2022.07.10.22277380

**Authors:** Tuomo Nieminen, Ilkka Kivekäs, Miia Artama, Hanna Nohynek, Jarno Kujansivu, Petteri Hovi

## Abstract

**Importance:** Spontaneous adverse reaction reports of sudden hearing loss have been observed and a population-based cohort study conducted in Israel showed an increase in the incidence of sudden sensorineural hearing loss (SSNHL) following vaccination with messenger RNA COVID-19 vaccine BNT162b2 (Pfizer-BioNTech). However, in this setting, possibility of confounding remained.

**Objective:** To assess a potential association between COVID-19 vaccinations and SSNHL.

**Design:** A register-based country-wide observational study with study period from January 1, 2019, until Apr 12, 2022.

**Setting:** Residents in Finland: 5,8 million.

**Participants:** All individuals identified from population information system alive or born during the study period except individuals having SSNHL during 2015 to 2018 according to specialized care derived diagnosis codes for SSNHL (ICD10-code H91.2) as a primary or secondary diagnosis.

**Exposures:** The *a priori* primary risk period was 0 to 54 days following each COVID-19 vaccine dose. The secondary risk time was 55 or more days after the vaccination. A later vaccine exposure overruled a previous one. A secondary analysis included a risk time of 0 to 54 days following a positive polymerase chain reaction (PCR) test for SARS-CoV-2.

**Main Outcome:** We compared incidences of SSNHL following COVID-19 vaccination to incidences before the COVID-19 epidemic in Finland. In our Poisson regression analysis, we adjusted for calendar time, age, sex, diabetes, cardiovascular disease, other chronic diseases, number of visits in primary health care.

**Results:** Comparison time constituted of 6.5 million person years, primary risk time of 1.7 million person years, and secondary risk time of 2.1 million person years. Before the epidemic yearly 18.7 / 100,000 people were diagnosed with SSNHL. Our data suggested no increased risk for SSNHL following any COVID-19 vaccination. In particular, adjusted incidence rate ratios, with 95 percent confidence intervals (95% CI) for the BNT162b2 vaccine’s three doses were 0.8 (95% CI 0.6 to 1.0), 0.9 (95% CI 0.6 to 1.2), and 1.3 (95% CI 0.9 to 2.0). SARS-CoV-2 infection was not associated with an increased incidence of SSNHL either.

**Conclusions and relevance:** Our results show no evidence of increased SSNHL with the COVID-19 vaccinations. We accounted for previous disease and other potential confounding factors. Our results base on diagnosis codes in specialized care but still need to be verified with settings, that are capable to evaluate the degree of hearing loss.

**Key Points:** *Question:* Are COVID-19 vaccinations associated with sudden sensorineural hearing loss (SSNHL), when assessed in a register-based country-wide observational study with data on potential confounding factors?

*Findings:* Our data suggested no increased risk for SSNHL following any COVID-19 vaccination.

*Meaning:* A large previous cohort study shows an increased risk for SSNHL following vaccine BNT162b2 (Pfizer-BioNTech) – our results, that take pre-existing disease into account, demonstrate no such an effect.

## Manuscript text

During the COVID-19 pandemic, there have been several case reports, where sudden sensorineural hearing loss (SSNHL) has been connected to the COVID-19 vaccination.^1,2^ The European database of suspected adverse drug reaction reports, as retrieved on March 15, 2022, includes more than 1,000 reports on COVID-19 9-vaccination related SSNHL.^3^ These reports have been rare, since the European authorities, in March, 2022, report the cumulative number of doses exceeded 1.5 billion doses.^4^ First national Vaccine Adverse Events Reporting System based larger data was published in 2021 from the USA.^5^ They concluded that the incidence of SSNHL was even lower after the COVID-19 vaccination than in non-vaccinated population. In the only population-based cohort study on this subject, from Israel, reported a statistically significant risk for SSNHL after BNT162b2 messenger RNA COVID-19 vaccine (Pfizer-BioNTech).^6^ The standardized incidence ratio was 1.4 when comparison was made to the year 2018 incidence in an age- and sex standardized population.

The association between SARS-CoV-2 infection and SSNHL is controversial. In a study by van Rijssen et al. there was no apparent relationship between SSNHL and COVID-19^7^. None of the 25 patients with SSNHL had a positive SARS-CoV-2 laboratory test result. However, in several case reports, SSNHL has followed SARS-CoV-2 infection.^8,9^

Studies that can account for the characteristics of the vaccinated are warranted. Here, we report such a study aiming at revealing a possible association between COVID-19 vaccinations and SSNHL. Secondarily, we investigate the associations between SARS-CoV-2 infection and SSNHL.

## Methods

In the population of 5.8 million residents in Finland, a COVID-19 vaccination campaign was initiated on December 26^th^, 2020. A priority was given to health care workers and to those with highest risk for COVID-19 related hospitalisation and death. This included the elderly, those with pre-existing diseases and those residing in nursing homes.^10^ Vaccination campaign was started with adenoviral vector vaccine ChAdOx1, but majority of the population was vaccinated with mRNA-vaccines. Of these, availability led to more immunizations with BNT162b2 than with mRNA.1273 with minuscule use of other vaccine products. Moreover, on October 7^th^, 2021, Finnish Institute for Health and Welfare (THL) recommended BNT162b2 for males under 30, due to data showing an increased risk of myocarditis in young men especially following 2^nd^ dose of mRNA.1273.^11^

We investigated a potential harmful effect of the COVID-19 vaccines by conducting an observational cohort study based on nationwide register data and studied the incidence of SSNHL in Finland. The study period was from January 1, 2019, until Apr 12, 2022 and the data was extracted from registers on Apr 27, 2022.

### Cohort

From the population information system, we identified all individuals alive or born during the study period. This register includes the birth date, gender, and unique personal identification code for all permanent residents of Finland. The personal identification code enables linkage to other registers, and we used it to retrieve exposure-, outcome-, and comorbidity information for the cohort. Individuals with specialised care diagnosis of sudden hearing loss, identified with the ICD-10 diagnosis code H91.2 from the Care Register for Health Care^12^ during 2015–2018, were removed from the cohort.

### Comorbidity data

We combined data from multiple sources to define pre-study comorbidity information for the cohort. Comorbidity conditions were identified until the start of the study period, December 31, 2018. The availability of information preceding 2018 depended on the data source: Starting from January 1, 2015, we included information from the Care register for health care^12^ and the Register for social assistance,^13^ and starting from January 1, 2018, from the register for reimbursement of medical expenses (Social Insurance Institute of Finland, SII).^14^ Based on these three data sources, dichotomic risk factors included in the analysis were diabetes, cardiovascular disease, nursing home residency, assisted living, and other institutional living. Other chronic diseases that increase risk for COVID-19 hospitalizations and mortality^15^ were included in the analyses as count (0,1,2,’3 or more’). Starting from January 1, 2015, the Register for primary health care visits^16^ provided a count (0,1,2, ‘3 or more’) of any primary care contacts as a proxy for care seeking behaviour. The included primary care contacts were individual visits, physical visits or remote visits, outpatient care, dental care, vaccinations, occupational health.

### Exposure data: vaccination and infection

The national vaccination register^17^ provided the vaccination dates and product names. Of vaccinations recorded within 14 days only the first date was utilized. We labelled each vaccination dose according to the most recent product.

To assess our secondary aim regarding SARS-CoV-2 infections we utilized data from the Finnish National Infectious Diseases Register.^18^ After January 1, 2020, we utilised the first positive SARS-CoV-2 laboratory test sample date per individual as the SARS-CoV-2 infection date. We ignored reoccurring infections. Most SARS-CoV-2 infections were detected based on a positive PCR laboratory test.

### Outcome data and follow-up

Diagnoses in specialized care visits and hospital ward periods are sent to the Care register for health care.^12^ For any individual, the first occurrence of the ICD-10 diagnosis code H91.2, ‘Sudden idiopathic hearing loss’, in the Care register for health care after January 1, 2019, was considered as an incident case of SSNHL. The follow-up of an individual ended at the time of the outcome or death or the end of the study, whichever happened first.

### Time-varying COVID-19 vaccination status

The vaccination status was defined as (i) pre-epidemic unvaccinated; (ii) epidemic unvaccinated; and (iii) vaccinated. The vaccination status for the entire cohort until February 29, 2020, was pre-epidemic unvaccinated, and then epidemic unvaccinated from March 1, 2020, until December 26, 2020, when the COVID-19 vaccinations in Finland started. For each vaccinated individual, the status first changed to pre-vaccination (30 days prior to the first vaccination) and then to vaccinated from the date of the first vaccination. The vaccinated state was further split into primary risk (0 to 54 days following any vaccination) and secondary risk (55 days and more following any vaccination) states, labelled by the dose and product of the latest vaccination. If individuals received multiple vaccine doses, a new primary risk period always overruled the previous period.

### Time varying SARS-CoV-2 infection status

The SARS-CoV-2 infection status was defined as uninfected or infected. The infection status for the whole cohort was first uninfected. Following the start of the COVID-19 epidemic the status of some changed to infected. The infected status was further split into primary and secondary risk periods, which were identical to those corresponding vaccination. Reinfections were not considered.

### Statistical analyses

We used a single Poisson regression model to estimate adjusted incidence rate ratios (aIRRs) between each vaccine exposure state and the pre-epidemic unvaccinated state and similarly between each infection exposure state and the uninfected state. Time-invariant covariates included in the regression were gender, chronic disease count, diabetes, cardiovascular disease, nursing home care, assisted living, other institutional living, and number of visits at primary care, as a proxy of care seeking behaviour. The time-dependent covariates were vaccination status, SARS-CoV-2 infection status, calendar month, and age group with lower group limits at ages 0, 12, 20, 30, 40, 50, 60, 70, and 80. To account for non-linear changes in the incidence of SSHNL by calendar month, we used a natural spline function.^19^ The number of knots used, 7, was chosen based on Akaike information criteria among choices 6 to 12.

We assumed that the incidence of SSHNL was piecewise constant depending on the time-dependent and time-invariant covariates. As all covariates were categorical, individuals contributed events and person time to follow-up groups based on the covariates. Each individual’s person time and number of SSHNL occurrences (zero or one) were then aggregated for all follow-up states.

Via Poisson regression we modelled the log incidence of SSNHL and estimated the regression parameters and their standard errors. We used a normal approximation to derive 95% confidence intervals (95% CI) for the parameter estimates on the log scale. Exponentiating the parameter estimates provided the adjusted incidence rate ratios (aIRR). We conducted the analyses with R version 4.2.^20^

### Sensitivity analyses

We learned two additional regression models to assess the impact of (i) calendar time adjustment, and (ii) risk factor and care-seeking adjustment, to the vaccination exposure aIRRs. The analyses were otherwise identical to our main analysis, but the first model excluded the calendar adjustment, and the second model further excluded the comorbidity and infection exposure covariates leaving only the following covariates: gender, age group and vaccination status.

### Ethics

A competent authority within the Finnish Institute for Health and Welfare declares further ethical review for the current research is not required. Surveillance including the safety of vaccines utilized in the national vaccination program is part of the Institute’s (THL) duties.^21^ THL has the statutory right, notwithstanding confidentiality provisions, to access and link necessary data from the national registers.

## Results

In the whole population of Finland between 2016 and 2019, the crude monthly incidence of SSNHL varied from 13 to 23 /100,000 person years (pyrs) (**Figure 1**). In 2020, April and May showed the lowest incidences after 2016 (11 and 12 / 100,000 pyrs) and February 2021 the highest incidence regarding this outcome (27 / 100,000 pyrs). Since then, the monthly incidences varied at the same level as before 2020, between 14 and 21 /100,000 pyrs.

**Figure 1.**
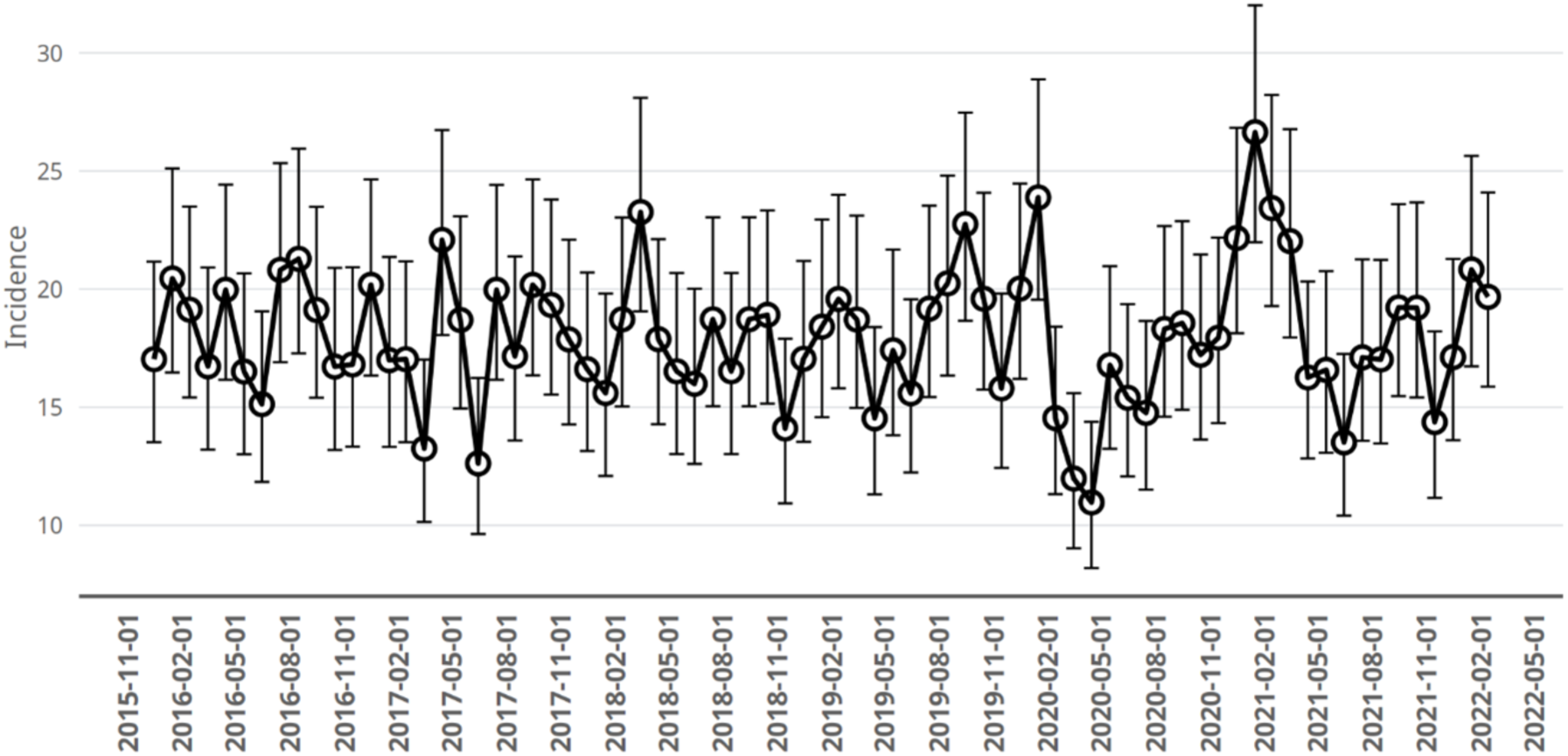
Incidence of sudden sensorineural hearing loss prior to and during the study period in Finland. Incidence per 100,000 person years of sudden sensorineural hearing loss in specialized care by calendar month. Vertical whiskers indicate 95 percent confidence intervals of the incidence, assuming a Poisson distribution of the monthly counts. The follow-up for the current study started January 1, 2019, and the epidemic in Finland started in March, 2020.

### Incidence during unvaccinated time

During follow-up time preceding the COVID-19 epidemic in Finland between January 1, 2019 and March 1, 2020, a total number of 1216 subjects in our cohort experienced SSNHL, within 6.5 million pyrs of follow-up time, and the crude incidence per 100, 000 pyrs was 18.7 (95% CI: 17.7 to 19.8) (**Table 1**). There was a sudden decrease in the incidence of SSNHL at the start of the epidemic in March 2020, followed by a slow increase back to the pre-epidemic level by the end of 2020 (**eFigure 1 in the Supplement**). During that time, the incidence of SSNHL was lower than before the epidemic, and was 15.7 (95% 14.6 to 16.8). Soon after the vaccinations began, during early 2021, the observed crude incidence during unvaccinated time decreased, likely due to the age- and health dependent vaccination campaign in Finland (**eFigure 1**).

**Table 1.**
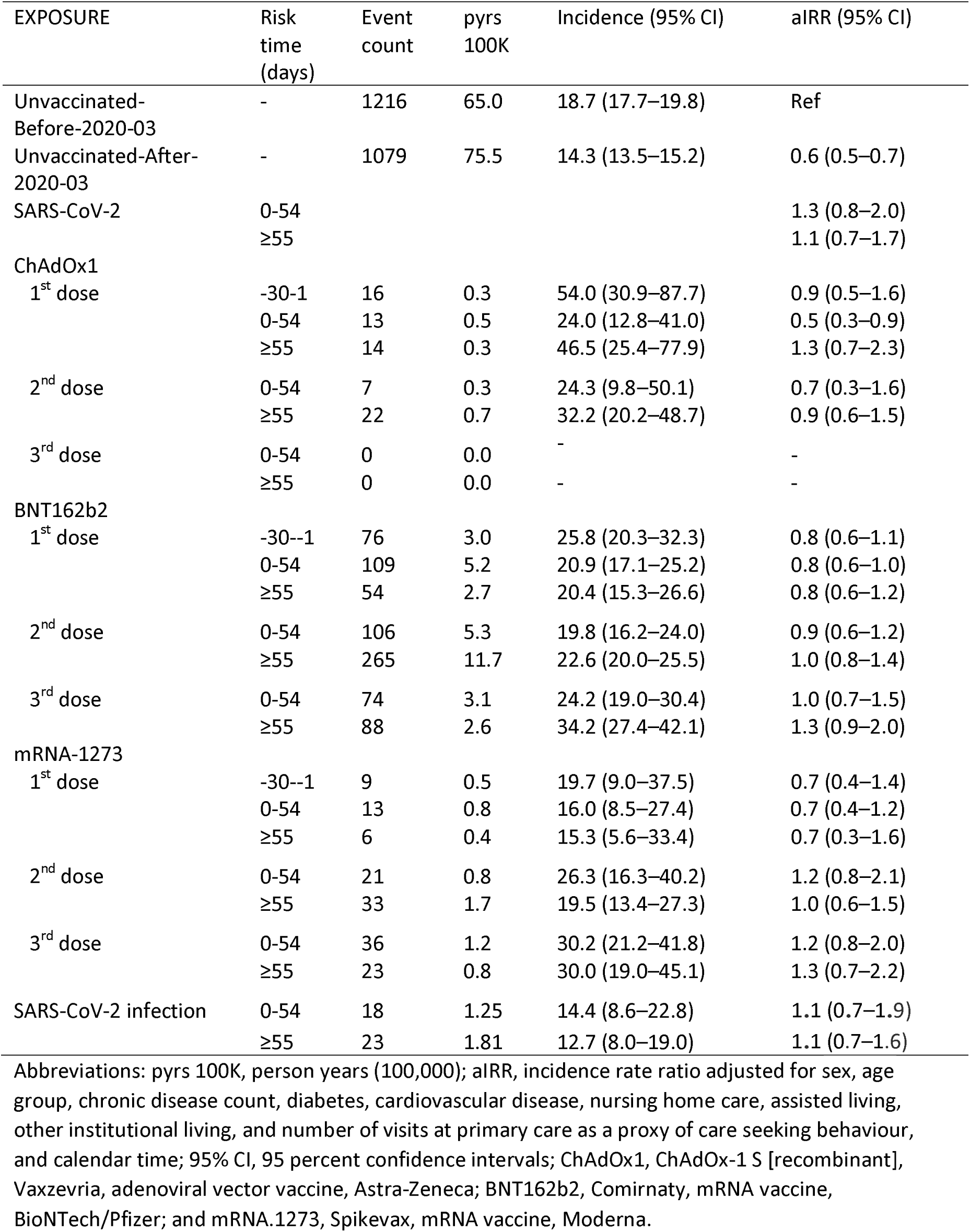
Incidences and adjusted incidence rate ratios of sudden sensorineural hearing loss.

| EXPOSURE | Risk time (days) | Event count | pyrs 100K | Incidence (95% CI) | aIRR (95% CI) |
| --- | --- | --- | --- | --- | --- |
| Unvaccinated- Before-2020-03 | - | 1216 | 65.0 | 18.7 (17.7–19.8) | Ref |
| Unvaccinated-After- 2020-03 | - | 1079 | 75.5 | 14.3 (13.5–15.2) | 0.6 (0.5–0.7) |
| SARS-CoV-2 | 0-54 |  |  |  | 1.3 (0.8–2.0) |
|  | ≥55 |  |  |  | 1.1 (0.7–1.7) |
| ChAdOx1 |  |  |  |  |  |
| 1<sup>st</sup> dose | -30-1 | 16 | 0.3 | 54.0 (30.9–87.7) | 0.9 (0.5–1.6) |
|  | 0-54 | 13 | 0.5 | 24.0 (12.8–41.0) | 0.5 (0.3–0.9) |
|  | ≥55 | 14 | 0.3 | 46.5 (25.4–77.9) | 1.3 (0.7–2.3) |
| 2<sup>nd</sup> dose | 0-54 | 7 | 0.3 | 24.3 (9.8–50.1) | 0.7 (0.3–1.6) |
|  | ≥55 | 22 | 0.7 | 32.2 (20.2–48.7) | 0.9 (0.6–1.5) |
| 3<sup>rd</sup> dose | 0-54 | 0 | 0.0 | - | - |
|  | ≥55 | 0 | 0.0 | - | - |
| BNT162b2 |  |  |  |  |  |
| 1<sup>st</sup> dose | -30--1 | 76 | 3.0 | 25.8 (20.3–32.3) | 0.8 (0.6–1.1) |
|  | 0-54 | 109 | 5.2 | 20.9 (17.1–25.2) | 0.8 (0.6–1.0) |
|  | ≥55 | 54 | 2.7 | 20.4 (15.3–26.6) | 0.8 (0.6–1.2) |
| 2<sup>nd</sup> dose | 0-54 | 106 | 5.3 | 19.8 (16.2–24.0) | 0.9 (0.6–1.2) |
|  | ≥55 | 265 | 11.7 | 22.6 (20.0–25.5) | 1.0 (0.8–1.4) |
| 3<sup>rd</sup> dose | 0-54 | 74 | 3.1 | 24.2 (19.0–30.4) | 1.0 (0.7–1.5) |
|  | ≥55 | 88 | 2.6 | 34.2 (27.4–42.1) | 1.3 (0.9–2.0) |
| mRNA-1273 |  |  |  |  |  |
| 1<sup>st</sup> dose | -30--1 | 9 | 0.5 | 19.7 (9.0–37.5) | 0.7 (0.4–1.4) |
|  | 0-54 | 13 | 0.8 | 16.0 (8.5–27.4) | 0.7 (0.4–1.2) |
|  | ≥55 | 6 | 0.4 | 15.3 (5.6–33.4) | 0.7 (0.3–1.6) |
| 2<sup>nd</sup> dose | 0-54 | 21 | 0.8 | 26.3 (16.3–40.2) | 1.2 (0.8–2.1) |
|  | ≥55 | 33 | 1.7 | 19.5 (13.4–27.3) | 1.0 (0.6–1.5) |
| 3<sup>rd</sup> dose | 0-54 | 36 | 1.2 | 30.2 (21.2–41.8) | 1.2 (0.8–2.0) |
|  | ≥55 | 23 | 0.8 | 30.0 (19.0–45.1) | 1.3 (0.7–2.2) |
| SARS-CoV-2 infection | 0-54 | 18 | 1.25 | 14.4 (8.6–22.8) | 1.1 (0.7–1.9) |
|  | ≥55 | 23 | 1.81 | 12.7 (8.0–19.0) | 1.1 (0.7–1.6) |

Abbreviations: pyrs 100K, person years (100,000); aIRR, incidence rate ratio adjusted for sex, age group, chronic disease count, diabetes, cardiovascular disease, nursing home care, assisted living, other institutional living, and number of visits at primary care as a proxy of care seeking behaviour, and calendar time; 95% CI, 95 percent confidence intervals; ChAdOx1, ChAdOx-1 S [recombinant], Vaxzevria, adenoviral vector vaccine, Astra-Zeneca; BNT162b2, Comirnaty, mRNA vaccine, BioNTech/Pfizer; and mRNA.1273, Spikevax, mRNA vaccine, Moderna.

### Incidence before and after vaccination

Within the COVID-19 vaccinated, the incidence of SSNHL did not show a temporal pattern related to vaccination, and the incidences during the main risk periods were similar to the incidences before vaccination and to those after the main risk periods (**Figure 2**). The crude incidences of SSNHL during the main risk period 0 to 54 days following the first COVID vaccine dose with ChAdOx1, BNT162b2, and mRNA.1273, were 24.0 (12.8 to 41.0), 20.9 (17.1 to 25.2), and 16.0 (8.5 to 27.4) per 100,000 pyrs and the aIRR estimates were less than one, indicating no increased risk of SSNHL after the first vaccination dose (**Table 1**). For the 30 days preceding the first vaccination doses, the aIRRs were similarly less than one for BNT162b2 and mRNA-1273, indicating no increase in the incidence from pre to post vaccination. For ChAdOx1 the aIRR estimate was lower during the main risk period than during the 30 days pre vaccination, and was 0.5 (0.3 to 0.9) versus 0.9 (0.5 to 1.6).

**Figure 2.**
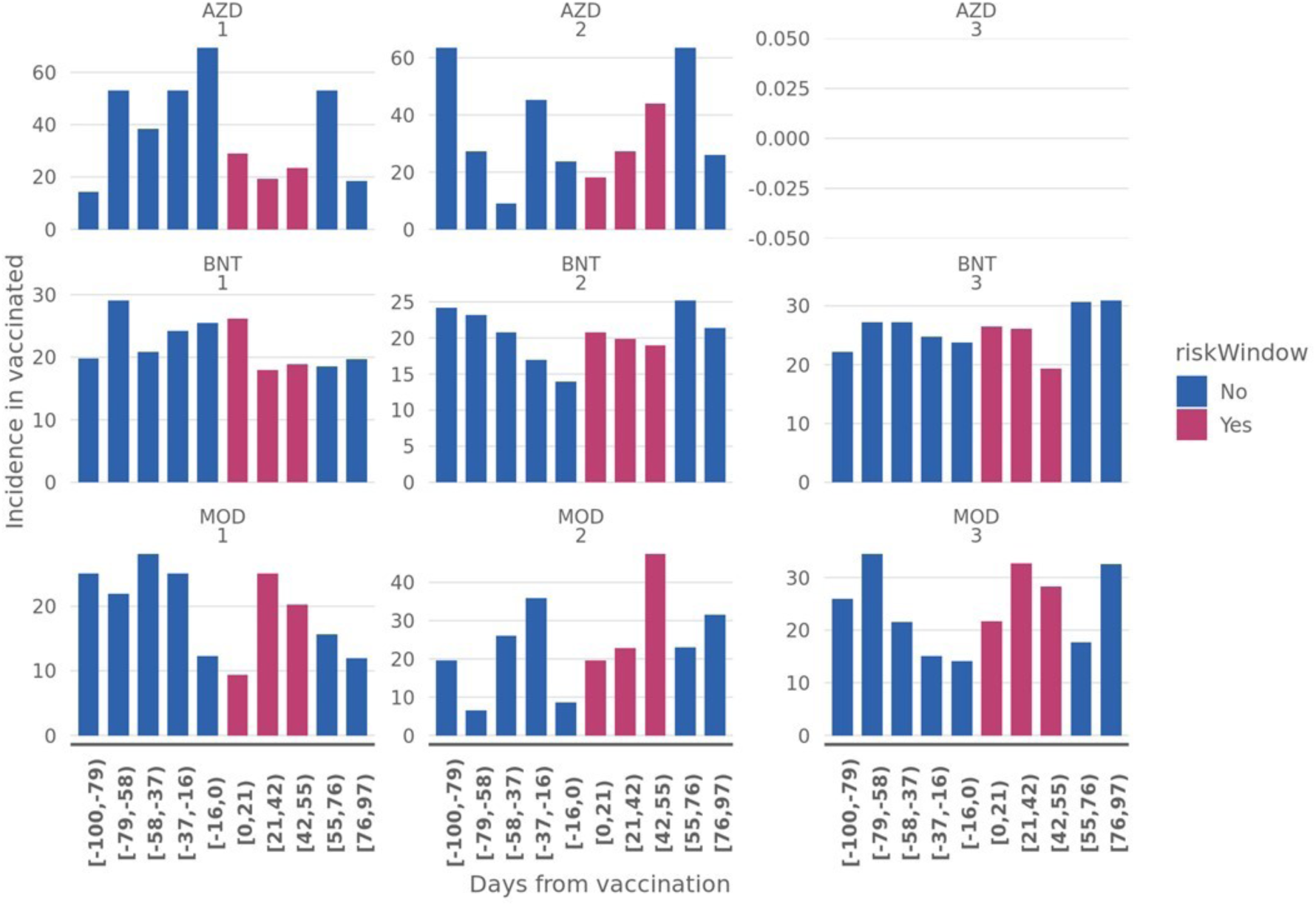
Incidence of sudden sensorineural hearing loss related to doses of COVID-19 vaccines. Incidence per 100,000 person years of sudden sensorineural hearing loss in specialized care by days from the vaccinations. Note that an individual case may be represented in adjacent panels, if interdose interval was less than 100 days. The primary risk time, 0 to 55 days from a vaccine dose is in purple, other time periods in blue. Numbers 1-3 indicate vaccine dose number. Abbreviations for vaccines: AZD: ChAdOx1, ChAdOx-1 S [recombinant], Vaxzevria, adenoviral vector vaccine, Astra-Zeneca; BNT: BNT162b2, Comirnaty, mRNA vaccine, BioNTech/Pfizer; MOD: mRNA.1273, Spikevax, mRNA vaccine Moderna.

Also the main and secondary risk periods following the second and third vaccinations showed no statistically significant differences to the pre-epidemic unvaccinated time in the adjusted analysis (**Table 1**).

### Incidence after infection

As our secondary aim, we explored how SARS-CoV-2 infection affects the risk for SSNHL. There was no evidence of an increased risk of SSNHL following SARS-CoV-2 infection. The aIRR for the main risk period 0 to 54 days following infection was 1.3 (95% CI: 0.8 to 2.0), and the aIRR for the secondary risk period 55 days onwards from infection was 1.1 (95% CI: 0.7 to 1.7), both indicating no significant change in the incidence of SSNHL when compared to the uninfected time.

### Sensitivity analyses

Removing the calendar adjustment produced similar or up to 25% higher vaccination-related aIRRs compared to the main analysis (**eTable 1 in the Supplement**). However, the aIRRs versus the pre-epidemic unvaccinated time were still mostly below one or close to one. The aIRR comparing the epidemic unvaccinated time to the pre-epidemic unvaccinated time was higher without calendar adjustment and was 0.9 (95% CI: 0.8–1.0). Removing the risk factor and care-seeking covariates from the model had almost no effect on the vaccination exposure results.

## Discussion

We investigated the proposed association between COVID-19 vaccinations and sudden sensorineural hearing loss (SSNHL) by conducting a nation-wide cohort study with high quality data sources. We compared the incidence of SSNHL following vaccinations to incidence before the COVID-19 epidemic in Finland. In order to remove possible confounding by age and sex we accounted for them in the analysis. Additionally, as opposed to previous studies, we also adjusted our analysis for individual differences in underlying diseases and care seeking behaviour, as well as temporal changes in the incidence of SSNHL unrelated to vaccination. In summary, we found no evidence of an impact of COVID-19 vaccination on SSNHL. Our adjusted incidence rate ratio (aIRR) estimates related to the main risk periods, from 0 to 54 days after vaccination, were, mostly close to one or even lower.

SSNHL is a rare, severe condition affecting roughly 5-20/100 000 people yearly in the developed world.^22,23^ In most cases the prognosis is good, and in a matter of weeks hearing will recover to normal level in 40-60 % of cases.^24, 25^ It is possible that a delay in or even absence of seeking help will decrease the incidence of diagnosed SSNHL cases. We found a reduction in the incidence of SSNHL during unvaccinated time during the COVID-19 epidemic, initiated by a sudden decrease in the incidence when the epidemic started. Reasons may include decrease in both demand and supply of health-care contacts, both leading to reduced encounters. During the COVID-pandemic such was shown by a reduced testing for autoimmune disease related antibodies.^26^ Regarding other possible reasons for lower incidence during the epidemic, we are unaware of any changes in the diagnosis processes or guidelines during the study, from 2019 to 2022.

One weakness of our study is that we defined SSNHL according to the diagnostic decision of each clinician and not to a standardized predefined definition. In a previous study by Härkönen et al.^25^ even 68% of SSNHL that were based only on diagnosis did not fulfil the criteria of SSNHL when audiograms were re-evaluated based on the American Academy of Otolaryngology– Head and Neck Surgery -guideline (sensorineural hearing loss of 30 dB or greater over at least three contiguous audiometric frequencies occurring within a 72-hr period).^27^ Audiograms, proper clinical history, and investigation were however most likely utilized, since we only used data on specialized where, according to our experience and internal data in THL, the vast majority of the cases are diagnosed by otorhinolaryngologists. Moreover, we have no reason to believe that any of the possible misclassification would be selectively occurring by vaccination status. Clearly, future research should be based on objective findings fulfilling international criteria for SSNHL and all included audiograms should be re-evaluated.^27^

One previous large-scale retrospective follow-up study has been recently conducted in Israel.^6^ In 21-day risk times following first and second dose BNT162b2 mRNA COVID-19 vaccine, small standardized incidence ratios were 1.4 and 1.2, and small attributable risks were 9 and 6 per one million vaccinees. Their comparison was based on incidence in the population during 2018 and 2019 and the analysis corrected for age and sex. However, for example, chronic disease, especially diabetes and cardiovascular disease remained as very likely confounders and the authors correctly conclude their results would need confirmation from elsewhere. Our results provided no confirmation for the Israeli study. This also is in line with a previous study from the USA, showing no association between SARS-CoV-2 vaccination and SSNHL.^5^

We defined a 55-day risk time following vaccination. A portion of our cases may have been initially evaluated in primary care with their first specialized care visit within a couple of weeks, and therefore we used a longer risk period compared to the previous Israeli study in which the risk period was 21 days following vaccination.^6^ It is notable that the Israeli study reported results from a secondary analysis utilising a 60-day risk period, with unchanged results. As we were unable to detect the exact symptom-onset dates from the registers and only were aware of the diagnosis dates, we may have misclassified a portion of cases to a risk time when their true classification would have been the preceding time period. However, only for misclassifications concerning dose 1 would this time-window have been unvaccinated time.

A peak in the SSNHL incidence around February 2021 (**Figure 1**). We are unaware of what causes this peak but it did not resemble the pattern seen in COVID-19 cases or COVID-19 deaths in the country.^28^ Also, in our analysis SARS-Cov-2 infection remained unassociated with any increased incidence of SSNHL and thus this peak was likely not due to SARS-CoV-2 infections. The vaccination dates were not clustered around the SSNHL peak either.^29^

We may have misclassified individuals as uninfected in case they were infected but did not seek health care or virus testing. Although virus testing is currently inadequate after the emergence of a fast-spreading Omicron virus variant, by early 2021 at least half of the SARS-COV-2 infections in the country had been detected and registered, according to results from serological surveys.^30^ That our large study revealed no connection between SARS-COV-2 infections and SSNHL parallels with a SSNHL patient series with a negative SARS-Co2 PCR tests.^7^ Other studies have reported isolated case reports where otherwise asymptomatic SSNHL patient have been tested with a positive SARS-Co2 PCR test^9^ and also highly symptomatic COVID-19 patients with concomitant SSNHL.^8^

In summary, our analysis accounted for individual patients’ characteristics including disease history and showed no evidence of the suspected association between COVID-19 vaccinations and SSNHL

## Conclusion

We set out to study, and possibly confirm, a signal of SSNHL following a messenger-RNA COVID-19 vaccine BNT162b2. We also investigated the possible signal following COVID-19 vaccines ChAdOx1 and mRNA.1273. We show no evidence of any effects by these vaccines on SSNHL. Studies with more detailed clinical data would be needed to reproduce our study with more objective diagnosis of sudden hearing loss.

## Supporting information

Letter from the head of department

## Data Availability

By Finnish law,the authors are not permitted to share individual-level register data. The computing code is available upon request. Possible inqueries.

## Acknowledgements

We thank Prof. Sangita Kulathinal, Department of Mathematics and Statistics, University of Helsinki, for her valuable help regarding the statistical methods. We also thank Esa Ruokokoski for contributions to data collection and management.

## Author’s contributions

I.K. literature search, study design, data collection, data interpretation, writing – original draft

T.N. conceptualisation, figures, data collection, study design, data analysis, data interpretation, writing – original draft, writing – review and editing, data curation, formal analysis

M.A. conceptualisation, literature search, study design, data analysis, data interpretation, writing – review & editing H.N. conceptualisation, literature search, study design, data interpretation, writing – review & editing, supervision

J.K. writing – review & editing

P.H. conceptualisation, literature search, study design, data analysis, data interpretation, writing – original draft, supervision

