## Supplement for "Sudden hearing loss following vaccination against COVID-19"

#### Supplementary Tables

##### eTable 1. All regression parameter estimates from the main analysis and from the two sensitivity analyses

| Covariate | aIRR (95% CI), main analysis | aIRR (95% CI), no calendar adjustment | aIRR (95% CI), no calendar, or comorbidity adjustment |
| --- | --- | --- | --- |
| (Intercept) | 8.02 (6.57–9.79) | 8.36 (7.03–9.93) | 14.29 (12.66–16.12) |
| Age (years) |  |  |  |
| 0-11 | 0.03 (0.01–0.05) | 0.03 (0.01–0.05) | 0.03 (0.01–0.05) |
| 12-19 | 0.23 (0.17–0.30) | 0.22 (0.17–0.30) | 0.24 (0.18–0.32) |
| 20-29 | 0.52 (0.43–0.63) | 0.52 (0.43–0.62) | 0.53 (0.44–0.64) |
| 30-39 | Ref | Ref | Ref |
| 40-49 | 1.36 (1.18–1.57) | 1.38 (1.20–1.59) | 1.39 (1.21–1.60) |
| 50-59 | 1.56 (1.37–1.79) | 1.59 (1.39–1.83) | 1.67 (1.46–1.91) |
| 60-69 | 1.85 (1.62–2.12) | 1.90 (1.66–2.17) | 2.14 (1.87–2.43) |
| 70-79 | 2.13 (1.85–2.45) | 2.21 (1.92–2.54) | 2.73 (2.40–3.11) |
| ≥80 | 1.55 (1.30–1.84) | 1.63 (1.38–1.93) | 1.95 (1.67–2.29) |
| Male gender (compared to female) | 1.12 (1.04–1.20) | 1.11 (1.04–1.19) | 1.09 (1.01–1.16) |
| Vaccine exposure (days from exposure) |  |  |  |
| Unvaccinated before March 2020 | Ref | Ref | Ref |
| Unvaccinated after March, 2020 | 0.57 (0.48–0.68) | 0.87 (0.81–0.95) | 0.87 (0.80–0.94) |
| ChAdOx1 |  |  |  |
| 1st dose (-30-1) | 0.90 (0.52–1.54) | 1.68 (1.02–2.76) | 1.77 (1.08–2.90) |
| (0-54) | 0.44 (0.24–0.82) | 0.69 (0.39–1.22) | 0.72 (0.41–1.28) |
| (≥55) | 1.23 (0.69–2.20) | 1.45 (0.85–2.45) | 1.53 (0.90–2.59) |
| 2nd dose (0-54) | 0.59 (0.25–1.38) | 0.60 (0.27–1.33) | 0.62 (0.28–1.38) |
| (≥55) | 0.94 (0.57–1.55) | 0.95 (0.63–1.45) | 0.98 (0.65–1.49) |
| 3rd dose (0-54) | 0.00 (0.00–Inf) | 0.00 (0.00–Inf) | 0.00 (0.00–Inf) |
| (≥55) | 0.00 (0.00–Inf) | 0.00 (0.00–Inf) | 0.00 (0.00–Inf) |
| BNT162b2 |  |  |  |
| 1st dose (-30-1) | 0.78 (0.57–1.07) | 1.24 (0.98–1.56) | 1.23 (0.98–1.55) |
| (0-54) | 0.77 (0.57–1.04) | 1.01 (0.83–1.23) | 1.00 (0.83–1.22) |
| (≥55) | 0.84 (0.58–1.21) | 0.92 (0.70–1.20) | 0.91 (0.69–1.19) |
| 2nd dose (0-54) | 0.84 (0.61–1.17) | 0.90 (0.73–1.09) | 0.89 (0.73–1.09) |
| (≥55) | 1.01 (0.74–1.38) | 1.02 (0.89–1.16) | 1.01 (0.89–1.16) |
| 3rd dose (0-54) | 0.97 (0.66–1.43) | 0.95 (0.75–1.19) | 0.95 (0.76–1.20) |
| (≥55) | 1.23 (0.82–1.84) | 1.07 (0.88–1.32) | 1.09 (0.89–1.33) |
| mRNA.1273 |  |  |  |
| 1st dose (-30-1) | 0.71 (0.36–1.42) | 1.06 (0.55–2.04) | 1.07 (0.55–2.05) |
| (0-54) | 0.78 (0.45–1.36) | 0.98 (0.59–1.62) | 0.98 (0.59–1.64) |
| (≥55) | 0.66 (0.29–1.53) | 0.70 (0.32–1.57) | 0.71 (0.32–1.59) |
| 2nd dose (0-54) | 1.16 (0.69–1.93) | 1.20 (0.77–1.87) | 1.21 (0.78–1.89) |
| (≥55) | 0.87 (0.55–1.36) | 0.86 (0.61–1.23) | 0.87 (0.62–1.24) |
| 3rd dose (0-54) | 1.15 (0.72–1.83) | 1.13 (0.81–1.59) | 1.13 (0.81–1.58) |
| (≥55) | 1.22 (0.73–2.03) | 1.04 (0.72–1.49) | 1.04 (0.72–1.50) |
| Calendar time adjustment with splines |  |  |  |
| parameter 1 | 2.81 (2.07–3.82) | - | - |
| parameter 2 | 0.83 (0.57–1.22) | - | - |
| parameter 3 | 1.41 (0.91–2.16) | - | - |
| parameter 4 | 0.86 (0.55–1.36) | - | - |
| parameter 5 | 1.22 (0.78–1.90) | - | - |
| parameter 6 | 0.98 (0.57–1.70) | - | - |
| parameter 7 | 0.74 (0.48–1.14) | - | - |
| SARS-CoV-2 infection |  |  |  |
| Uninfected time | Ref | Ref |  |
| (0-54) | 1.17 (0.74–1.86) | 1.09 (0.69–1.72) | - |
| (≥55) | 1.07 (0.70–1.63) | 0.93 (0.62–1.41) | - |
| Risk factors (Yes vs No) |  |  |  |
| Diabetes (any type) | 0.99 (0.89–1.10) | 0.99 (0.89–1.10) | - |
| Cardiovascular disease | 1.22 (1.12–1.34) | 1.22 (1.12–1.34) | - |
| Assisted living | 0.14 (0.06–0.32) | 0.14 (0.06–0.32) | - |
| Elderly home | 0.28 (0.07–1.13) | 0.28 (0.07–1.13) | - |
| Institutional living | 0.38 (0.10–1.53) | 0.38 (0.09–1.52) | - |
| Number of other risk factors |  |  |  |
| 0 | Ref | Ref |  |
| 1 | 1.17 (1.06–1.29) | 1.17 (1.07–1.29) | - |
| 2 | 1.28 (1.02–1.61) | 1.28 (1.02–1.62) | - |
| ≥3 | 2.09 (1.18–3.69) | 2.09 (1.19–3.70) | - |
| Number of primary health care contacts |  |  |  |
| 0 | Ref | Ref |  |
| 1 | 1.49 (1.20–1.85) | 1.50 (1.21–1.86) | - |
| 2 | 1.61 (1.29–1.99) | 1.62 (1.30–2.01) | - |
| ≥3 | 1.85 (1.61–2.12) | 1.86 (1.62–2.14) | - |

Abbreviations: aIRR adjusted incidence rate ratio; 95% CI, 95 percent confidence interval; Ref, referent group; ChAdOx1: ChAdOx-1 S [recombinant], Vaxzevria, adenoviral vector vaccine, Astra-Zeneca; BNT162b2: Comirnaty, mRNA vaccine, BioNTech/Pfizer; mRNA.1273: Spikevax, mRNA vaccine Moderna. Primary care contacts were retrieved from the Register for primary health care visits. The following conditions were required for the contact to be included: individual visits, physical visits or remote visits, outpatient care, dental care, vaccinations, occupational health. The other risk factors included the following conditions: Chronic kidney disease, Lung disease, Chronic liver disease, Neurological Disorders, Addison’s disease, Schizophrenia, Down syndrome, Immunosuppressive disease/medication.

Supplementary figures

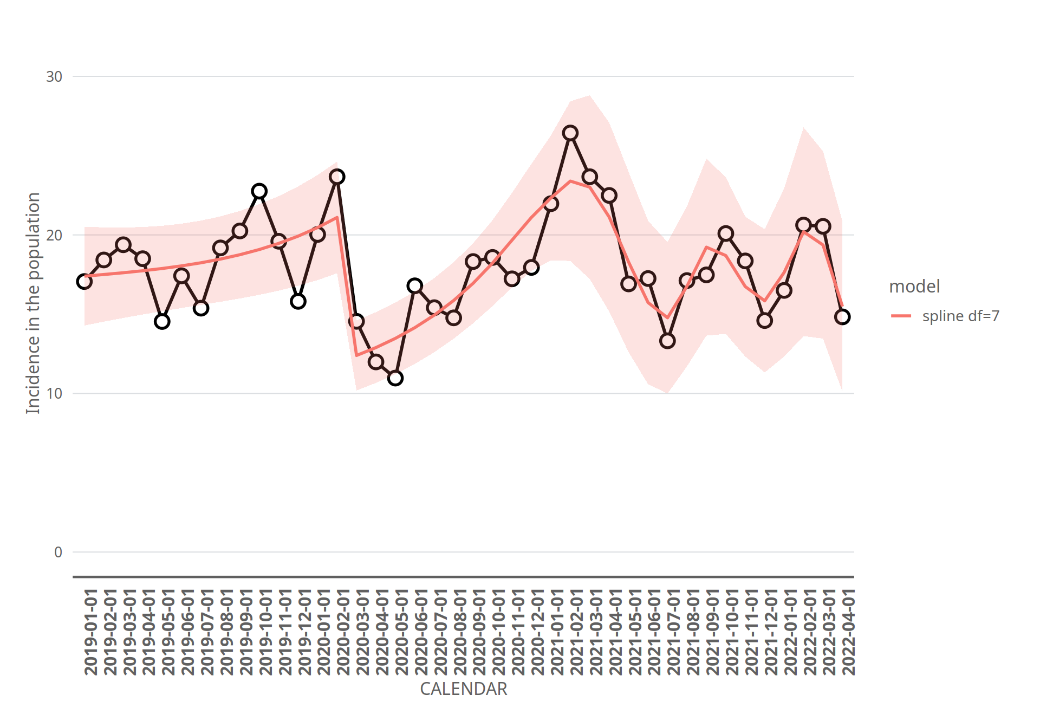

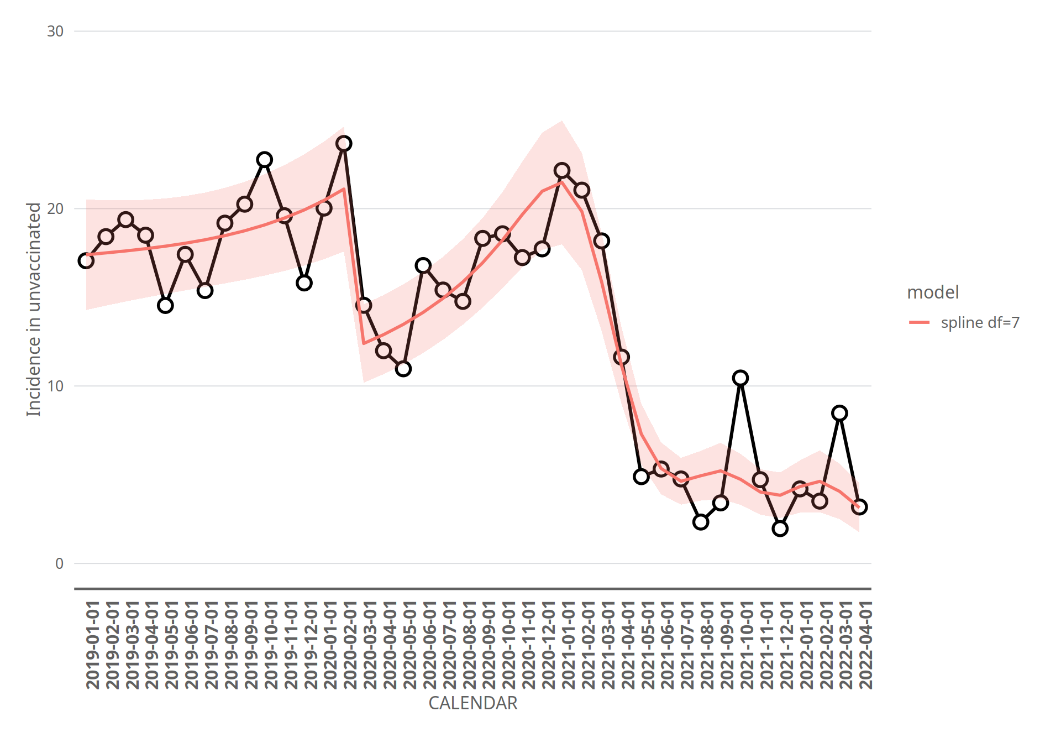

eFigure 1: Incidence of sudden sensorineural hearing loss during the study period.

Incidence per 100,000 person years of sudden sensorineural hearing loss in specialized care by calendar month. The follow- up for the current study started January 1, 2019, and the COVID-19 epidemic in Finland started March, 2020. Black dots and black line represent the observed incidence. The red line and area represent predictions from our main model, and the shaded areas correspond to pointwise 95% confidence intervals. The model takes into account calendar time, background variables such as gender, age and previous diseases, as well as the time-dependent vaccination status, where the unvaccinated time is split into pre-epidemic and epidemic. Upper panel: Incidence for the whole population. There was a sudden drop in the incidence when the epidemic started, which is captured by our model via the change of the cohort from pre-epidemic unvaccinated time into the epidemic unvaccinated time. Of note is that the adjusted incidence rare ratio estimate between epidemic unvaccinated time and pre-epidemic unvaccinated time in our main model mostly describes this sudden change in incidence. Lower panel: Incidence during unvaccinated time. The incidence during unvaccinated time decreased after the COVID-19 vaccination program started during the beginning of 2021. This can be explained by age- and health related selection into the COVID-19 vaccination program, leaving younger and healthier individuals into the unvaccinated group.
