## Supplementary material for "Sudden hearing loss following vaccination against COVID-19": Letter from the head of department

thl

To whom it may concern,

As director of the department for Health security of the Finnish Institute for Health and Welfare, I certify that:

- I am the competent auhority for assessing whether research requires institutional ethical review or if the Finnish Communicable Diseases Act (Tartuntatautilaki 1227/2016, https://www.finlex.fi/en/laki/kaannokset/2016/en20161227) and the law on the duties of the Finnish Institute for Health and Welfare (Laki Terveyden ja hyvinvoinnin laitoksesta 668/2008, https://www.finlex.fi/en/laki/kaannokset/2008/en20080668_20080668.pdf) allow the implementation of the research without seeking further ethical review.

- The research presented by **Nieminen T** et al. in **“Sudden hearing loss following vaccination against COVID-19”** did not require further ethical review before implementation as its aim was to monitor vaccine safety (Tartuntatautilaki 1227/2016).

Helsinki, July 8, 2022

Prof Mika Salminen
